# Parenting Self-Efficacy within the First Year Postpartum in Non-Birthing Parents: A Scoping Review Protocol

**DOI:** 10.1101/2023.10.19.23297279

**Authors:** Justine Dol, Jennifer A. Parker, Christine T Chambers, Phillip Joy, Patricia Leahy-Warren, Cindy-Lee Dennis, Marilyn Macdonald, Kristy Hancock

## Abstract

**Objective:** The objective of this scoping review is to identify and analyze the literature on parenting self-efficacy in the non-birthing parent within the first year postpartum.

**Introduction:** Parenting self-efficacy is a parent’s confidence in their ability to organize and execute a set of tasks related raising children and providing them with protection and care in order to ensure their healthy development. To date, much of the work on parenting selfefficacy has focused on the birthing parent (e.g., mothers) excluding the non-birthing parent (e.g., fathers and LGBTQ+ parents).

**Inclusion criteria:** Studies that include participants that identify as the non-birthing parent (regardless of sex or gender) who have had an infant less than 12 months of age. Various self-efficacy outcomes related to parenting will be included, as well as sub-types of selfefficacy (e.g., chest/breastfeeding). No limitations based on context, geography, or parity will be applied. All qualitative and quantitative study designs will be included.

**Methods:** Databases to be searched will include MEDLINE, Embase, Web of Science, CINAHL, and PsycINFO with no time limitations applied. Title and abstracts and full text will be screened by two reviewers with disagreements solved with a third reviewer. The data extracted will include specific details about the population, study, methods, and outcomes. Data will be reported narratively and in tables, as appropriate.

## Introduction

Parenting self-efficacy has been defined as “beliefs or judgments a parent holds of their capabilities to organize and execute a set of tasks related to parenting a child” (p.390).^1^ Bandura, the pioneer of self-efficacy theory, suggested that the greater a person’s confidence related to a specific behaviour (i.e., parenting), the more likely they are to engage in that behaviour.^2^ Thus, the development of parenting self-efficacy is an essential component in the establishment of high-quality parenting habits that promote optimal child development.

Parenting self-efficacy can be examined targeting either general self-efficacy across the parenting role or task specific self-efficacy (e.g., breastfeeding).^3^ In a systematic review on self-report measures of parenting self-efficacy, Wittkowski et al.^3^ identified 34 different measures, none of which have been consistently used in the literature. Furthermore, none have been specifically developed for the non-birthing parent.^4^ There is also variation in the level of self-efficacy measurement, which can include global (beliefs about an ability to complete any task), domain-general (beliefs about being able to complete a specific task – i.e., parenting), and domain-specific (beliefs about being able to complete a specific task in certain area or conditions – i.e., breastfeeding, newborn stage).^2,4^ While global self-efficacy is not appliable in the current review, both domain-general and domain-specific can be related to parenting and parenting behaviours.

To date, much of the parenting self-efficacy work has focused on the birthing person or mother. Maternal self-efficacy has been found to positively influence parenting behaviour, which is associated with positive newborn and child outcomes.^5,6^ Various factors have been found to influence feelings of maternal self-efficacy, including parity,^7,8^ levels of social support,^9,10^ and postpartum anxiety and depression.^11,12^

Overall, there has been a general lack of parenting self-efficacy research on the non-birthing parent and the work completed thus far is disjointed. Previous work comparing mothers and fathers’ found that mothers had greater parenting self-efficacy than fathers^13^ while others have found no difference.^14^ Importantly, paternal self-efficacy has been found to be positively associated with supportive, early, and engaged parenting,^15,16^ suggesting that fathers’ level of self-efficacy is important for child and family outcomes. Correlates of greater paternal parenting self-efficacy have been found to contribute to a positive coparenting relationship,^17^ increased social support,^14^ enhanced parenting satisfaction,^14^ and lower parenting stress.^18,19^ What is currently lacking is a collation and summarization of all literature to date to guide future work.

There is even less research on parenting self-efficacy in non-heterosexual parenting relationships, including various configurations of LGBTQ+ partners.^20,21^ This is despite a growing number of non-heterosexual family structures, with the percentage of same-sex child-rearing couples in Canada growing from 8.6% in 2001 to 12.0% in 2016.^22^ In Canada, 23.6% of the heterosexual population were considered parents with a child under the age of 18, compared to 13.4% of bisexual people and 6.2% of gay and lesbian individuals.^23^ In the United States, approximately 19% of same-sex couple households had a child under the age of 18.^24^ In one study that compared heterosexual-parents, gay-father families, and lesbian mother-families no differences between family type on perceptions of parental self-efficacy.^25^ Conversely, another study found that lesbian mothers, heterosexual mothers, and gay fathers all reported higher parenting self-efficacy than heterosexual fathers. As the number of nonheterosexual parents continue to grow, it is important to synthesize the current evidence related to parenting self-efficacy in the non-birthing parent and to compare differences among various populations (e.g., gay fathers, lesbian mothers, transgendered parents, heterosexual fathers)

A preliminary search of MEDLINE, the Cochrane Database of Systematic Reviews and JBI Evidence Synthesis was conducted and no current or underway systematic reviews or scoping reviews on the topic were identified. The objective of this scoping review is to identify and analyze the literature on parenting self-efficacy in the non-birthing parents within the first year postpartum.

### Eligibility Criteria

#### Participants

The review will consider studies that include participants that identify as the non-birthing parent (regardless of sex or gender). Parenting self-efficacy will span the whole postpartum period – from birth to 12 months post-birth. Both biological and adoptative parents will be included, regardless of conception method (e.g., natural, surrogacy, in vitro fertilization). No geographical or parity limitations will be applied. Parents of both full-term and preterm infants will be included as well as singletons and multiples. Studies that solely reported on maternal outcomes or did not separate out paternal outcomes (e.g., reports on parents broadly) will be excluded. Studies that report across the perinatal period, including the antenatal period, will be included.

#### Concept

This review will seek to map the data on parenting self-efficacy outcomes relevant to the non-birthing parent (regardless of sex or gender) in their role as a parent as reported in the literature. Studies reporting on domain-general (beliefs about being able to complete a specific task – i.e., parenting self-efficacy) or domain-specific (beliefs about being able to complete a specific task in certain area or conditions – i.e., breastfeeding self-efficacy) will be included.

It is important to note that there is significant variation in terminology used to discuss parenting self-efficacy. For instance, in the literature, parenting confidence, parenting selfefficacy, and perceived competence have been used interchangeably with inconsistent conceptual definitions.^4^ Given that parenting self-efficacy is the most consistently used term,^4^ this will be used throughout the review, with the understanding that some studies may individually use the term competence and confidence. As such, these terms will be included in the search strategy for this review in order to capture all relevant studies.

#### Context

Existing scoping reviews have primarily focused on qualitative studies looking at first time fathers across the perinatal period ^26^ or included all first-time parents ^27^. Thus, there is a gap in evidence synthesis on the experience of the non-birthing parent’s parenting selfefficacy and those who are multiparous.

#### Type of Sources

This scoping review will consider both experimental and quasi-experimental study designs including randomized controlled trials, non-randomized controlled trials, before and after studies and interrupted time-series studies. In addition, analytical observational studies including prospective and retrospective cohort studies, case-control studies and analytical cross-sectional studies will be considered for inclusion. This review will also consider descriptive observational study designs including case series, individual case reports and descriptive cross-sectional studies for inclusion.

Further, qualitative studies will be considered that include, but are not limited to, designs such as phenomenology, grounded theory, ethnography, qualitative description, action research and feminist research.

Mixed method studies will also be included. Reviews (systematic, non-systematic, scoping, etc.) will be excluded but will be evaluated for original studies to be included, if applicable. Letters to the editor, editorials, commentaries, conference abstracts, dissertations, books, book chapters, and grey literature will not be included. If less than five peer reviewed studies are available for analysis, preprints will be searched for additional studies. Otherwise, preprints will not be included as they have not yet been peer-reviewed.

## Method

This review will be reported following the PRISMA extension for Scoping Reviews^28^ and will be conducted in accordance with the Joanna Briggs Institute (JBI) methodology for scoping review.^29^

### Search strategy

The search strategy was developed in consultation with a health science librarian and will aim to locate published studies. An initial limited search of MEDLINE was undertaken to identify articles on the topic. The text words contained in the titles and abstracts of relevant articles, and the index terms used to describe the articles were used to develop a full search strategy for PSYCInfo (see Appendix 1). The search strategy was peer-reviewed by a second health sciences librarian using the Peer Review of Electronic Search Strategies (PRESS) Guideline^30^ and refined according to the feedback received. The final search strategy, including all identified keywords and index terms, will be adapted for each included database and/or information source. Databases to be searched include MEDLINE All (Ovid), Embase (Elsevier), CINAHL Full Text (EBSCO), Scopus (Elsevier), and PsycINFO (EBSCO). The reference list of all included sources of evidence will be screened for additional studies using Citation Chaser or Scopus. No time limitations will be applied.

### Study selection

All identified citations through the search will be uploaded to Covidence^31^ and duplicates will be removed through their automation process. After a pilot of inclusion/exclusion criteria, title and abstracts and full text will be screened by two reviewers with disagreements solved with a third reviewer or discussion. Reasons for exclusion at the full-text stage will be reported. Results of the search will be reported in full in the final scoping review and presented in a PRISMA flow diagram.^28^

### Data extraction & synthesis

The data extracted will include specific details about the population, study, methods, and outcomes. Data extraction will be piloted before full data are extracted by one reviewer and verified by another. The findings will be presented in narrative form including tables and figures, where appropriate. For qualitative studies, data related to primary themes will be extracted. For quantitative studies, data related to rates of self-efficacy, relation to other populations (e.g., mothers), measurement tools, and correlated outcomes (e.g., stress, anxiety, depression) will be extracted as available and appropriate. Consideration for population characterises will be considered in the analysis (e.g., comparing parenting self-efficacy in parents of full-term and pre-term infants).

## Data Availability

Not applicable - protocol.

## Appendix I Search strategy for APA PsycInfo

Appendix I: Search strategy for APA PsycInfo
| # | Query | Limiters/Expanders |
| --- | --- | --- |
| S1 | TI ( "self-efficacy" or confiden* or competen* or agency ) OR AB ( "self-efficacy" or confiden* or competen* or agency ) | Expanders - Apply related words;<br>Apply equivalent subjects<br>Search modes - Boolean/Phrase |
| S2 | DE "Self-Efficacy" | Expanders - Apply related words;<br>Apply equivalent subjects<br>Search modes - Boolean/Phrase |
| S3 | DE "Self-Confidence" | Expanders - Apply related words;<br>Apply equivalent subjects<br>Search modes - Boolean/Phrase |
| S4 | DE "Competence" | Expanders - Apply related words;<br>Apply equivalent subjects<br>Search modes - Boolean/Phrase |
| S5 | DE "Agency" | Expanders - Apply related words;<br>Apply equivalent subjects<br>Search modes - Boolean/Phrase |
| S6 | S1 OR S2 OR S3 OR S4 OR S5 | Expanders - Apply related words;<br>Apply equivalent subjects<br>Search modes - Boolean/Phrase |
| S7 | TI ( postpartum or "post-partum" or postnatal or "post-natal" or perinatal or "peri-natal" or "post-birth" or neonat* or infant* or newborn# or baby or babies ) OR AB ( postpartum or "post-partum" or postnatal or "post-natal" or perinatal or "peri-natal" or "post-birth" or neonat* or infant* or newborn# or baby or babies ) | Expanders - Apply related words;<br>Apply equivalent subjects<br>Search modes - Boolean/Phrase |
| S8 | DE "Postnatal Period" | Expanders - Apply related words;<br>Apply equivalent subjects<br>Search modes - Boolean/Phrase |
| S9 | DE "Neonatal Period" | Expanders - Apply related words;<br>Apply equivalent subjects<br>Search modes - Boolean/Phrase |
| S10 | S7 OR S8 OR S9 | Expanders - Apply related words;<br>Apply equivalent subjects<br>Search modes - Boolean/Phrase |
| S11 | TI ( father* or paternal or dad or dads or stepparent* or "step-parent*" or stepfather* or "step-father*" or stepdad# or "step-dad#" or coparent* or "co-parent*" or nonbirthing or "non-birthing" ) OR AB ( father* or paternal or dad or dads or stepparent* or "step-parent*" or stepfather* or "step-father*" or stepdad# or "step-dad#" or | Expanders - Apply related words;<br>Apply equivalent subjects<br>Search modes - Boolean/Phrase |
|  | coparent* or "co-parent*" or nonbirthing or "non-birthing" ) |  |
| S12 | TI ( (agender or androgynous or bicurious or bigender or bisexual* or "cross sex" or crossgender or gay or "gender fluid" or "gender non-conform*" or genderfluid or genderfuck* or genderless or genderqueer or GLB* or heteroflexible or homosexual* or intersex or lesbian* or lesbigay or LGB* or nonbinary or "non-binary" or nongendered or "non-gendered" or queer or "same gender" or "same sex" or "third gender" or trans* or "two-spirit") N1 (parent* or mother* or maternal) ) OR AB ( (agender or androgynous or bicurious or bigender or bisexual* or "cross sex" or crossgender or gay or "gender fluid" or "gender non-conform*" or genderfluid or genderfuck* or genderless or genderqueer or GLB* or heteroflexible or homosexual* or intersex or lesbian* or lesbigay or LGB* or nonbinary or "non-binary" or nongendered or "non-gendered" or queer or "same gender" or "same sex" or "third gender" or trans* or "two-spirit") N1 (parent* or mother* or maternal) ) | Expanders - Apply related words;<br>Apply equivalent subjects<br>Search modes - Boolean/Phrase |
| S13 | TI ( (adopt* or foster*) N1 (parent* or mother* or maternal) ) OR AB ( (adopt* or foster*) N1 (parent* or mother* or maternal) ) | Expanders - Apply related words;<br>Apply equivalent subjects<br>Search modes - Boolean/Phrase |
| S14 | DE "Adoptive Parents" OR DE "Fathers" OR DE "Foster Parents" OR DE "Homosexual Parents" OR DE "Stepparents" | Expanders - Apply related words;<br>Apply equivalent subjects<br>Search modes - Boolean/Phrase |
| S15 | S11 OR S12 OR S13 OR S14 | Expanders - Apply related words;<br>Apply equivalent subjects<br>Search modes - Boolean/Phrase |
| S16 | S6 AND S10 AND S15 | Limiters - Publication Type: All Journals<br>Expanders - Apply related words;<br>Apply equivalent subjects<br>Search modes - Boolean/Phrase |

## References

1. de Montigny F, Lacharite Carl. Perceived parental efficacy: Concept analysis. J Adv Nurs. 2005;49(4):387–96.

2. Bandura A. Self-efficacy: Toward a unifying theory of behavioral change. Psychol Rev. 1977;84(2):191–215.

3. Wittkowski A, Garrett C, Calam R, Weisberg D. Self-Report Measures of Parental Self-Efficacy: A Systematic Review of the Current Literature. J Child Fam Stud. 2017;26(11):2960–78.

4. Vance AJ, Brandon DH. Delineating among Parenting Confidence, Parenting Self-Efficacy and Competence. ANS Adv Nurs Sci. 2017;40(4):E18–37.

5. Jones TL, Prinz RJ. Potential roles of parental self-efficacy in parent and child adjustment: A review. Clin Psychol Rev. 2005;25(3):341–63.

6. Shorey S, Chan SWC, Chong YS, He HG. Maternal parental self-efficacy in newborn care and social support needs in Singapore: A correlational study. J Clin Nurs. 2014;23(15–16):2272–83.

7. Bryanton J, Gagnon AJ, Hatem M, Johnston C. Predictors of early parenting selfefficacy: Results of a prospective cohort study. Nurs Res. 2008;57(4):252–9.

8. Dol J, Richardson B, Grant A, Aston M, McMillan D, Tomblin Murphy G, et al. Influence of parity and infant age on maternal self-efficacy, social support, postpartum anxiety, and postpartum depression in the first six months in the Maritime Provinces, Canada. Birth. 2021;48(3):438–47.

9. Haslam D, Pakenham K, Smith A. Social support and postpartum depressive symptomatology: The mediating role of maternal self-efficacy. Infant Ment Health J. 2006;27(3):276–91.

10. Esmaelzadeh Saeieh S, Rahimzadeh M, Yazdkhasti M, Torkashvand S. Perceived social support and maternal competence in primipara women during pregnancy and after childbirth. Int J Community Based Nurs Midwifery. 2017;5(4):408–16.

11. Kohlhoff J, Barnett B. Parenting self-efficacy: Links with maternal depression, infant behaviour and adult attachment. Early Hum Dev. 2013;89(4):249–56.

12. Reck C, Noe D, Gerstenlauer J, Stehle E. Effects of postpartum anxiety disorders and depression on maternal self-confidence. Infant Behav Dev. 2012;35(2):264–72.

13. Brage Hudson D, Elek SM, Ofe Fleck M. FIRST-TIME MOTHERS’ AND FATHERS’ TRANSITION TO PARENTHOOD: Infant Care Self-Efficacy, Parenting Satisfaction, and Infant Sex. Issues Compr Pediatr Nurs. 2001;24(1):31–43.

14. Yang X, Ke S, Gao LL. Social support, parental role competence and satisfaction among Chinese mothers and fathers in the early postpartum period: A cross-sectional study. Women Birth J Aust Coll Midwives. 2020 May;33(3):e280–5.

15. Murdock K. An Examination of Parental Self-Efficacy Among Mothers and Fathers. Psychol Men Masculinity. 2013 Feb 7;14:314–23.

16. Shorey S, Ang L, Goh E, Gandhi M. Factors influencing paternal involvement during infancy: A prospective longitudinal study. J Adv Nurs. 2018;75.

17. Donithen R, Schoppe-Sullivan S. Correlates and predictors of parenting self-efficacy in new fathers. J Fam Psychol. 2022 Apr;36(3):396–405.

18. Fang Y, Boelens M, Windhorst DA, Raat H, van Grieken A. Factors associated with parenting self-efficacy: A systematic review. J Adv Nurs. 2021;77(6):2641–61.

19. Gordo L, Oliver-Roig A, Martínez-Pampliega A, Elejalde LI, Fernández-Alcantara M, Richart-Martínez M. Parental perception of child vulnerability and parental competence: The role of postnatal depression and parental stress in fathers and mothers. PLOS ONE. 2018 Aug 27;13(8):e0202894.

20. Hafford-Letchfield T, Cocker C, Rutter D, Tinarwo M, McCormack K, Manning R. What do we know about transgender parenting?: Findings from a systematic review. Health Soc Care Community. 2019;27(5):1111–25.

21. Biocco R, Carone N, Ioverno S, Lingiardi V. Same-Sex and Different-Sex Parent Families in Italy: Is Parents’ Sexual Orientation Associated with Child Health Outcomes and Parental Dimensions? J Dev Behav Pediatr JDBP. 2018;39(1):555–63.

22. Government of Canada SC. Census in Brief: Same-sex couples in Canada in 2016 [Internet]. 2017 [cited 2023 Aug 15]. Available from: https://www12.statcan.gc.ca/census-recensement/2016/as-sa/98-200-x/2016007/98-200-x2016007-eng.cfm

23. Government of Canada SC. Family and household characteristics of lesbian, gay and bisexual people in Canada [Internet]. 2021 [cited 2023 Aug 15]. Available from: https://www150.statcan.gc.ca/n1/pub/89-28-0001/2018001/article/00021-eng.htm

24. Gates G. LGBT Parenting in the United States [Internet]. Williams Institute. [cited 2023 Aug 15]. Available from: https://williamsinstitute.law.ucla.edu/publications/lgbt-parenting-us/

25. Rubio B, Vecho O, Gross M, van Rijn-van Gelderen L, Bos H, Ellis-Davies K, et al. Transition to parenthood and quality of parenting among gay, lesbian and heterosexual couples who conceived through assisted reproduction. J Fam Stud. 2020 Aug;26(3):422–40.

26. Leahy-Warren P, Philpott L, Elmir R, Schmied V. Fathers’ perceptions and experiences of support to be a parenting partner during the perinatal period: A scoping review. J Clin Nurs. 2023;32(13–14):3378–96.

27. Sæther KM, Berg RC, Fagerlund BH, Glavin K, Jøranson N. First-time parents’ experiences related to parental self-efficacy: A scoping review. Res Nurs Health. 2023;46(1):101–12.

28. Tricco AC, Lillie E, Zarin W, O’Brien KK, Colquhoun H, Levac D, et al. PRISMA extension for scoping reviews (PRISMA-ScR): Checklist and explanation. Ann Intern Med. 2018;169(7):467–73.

29. Joanna Briggs Institute. The Joanna Briggs Institute reviewers’ manual 2015 methodology for JBI scoping reviews. South Australia; 2015.

30. McGowan J, Sampson M, Salzwedel DM, Cogo E, Foerster V, Lefebvre C. PRESS Peer Review of Electronic Search Strategies: 2015 Guideline Statement. J Clin Epidemiol. 2016 Jul;75:40–6.

31. Veritas Health Innovation. Covidence Systematic Review Software [Internet]. Melbourne, Australia; Available from: http://www.covidence.org

